# A Bayesian estimate of the COVID-19 infection fatality ratio in Brazil based on a random seroprevalence survey

**DOI:** 10.1101/2020.08.18.20177626

**Authors:** Valerio Marra, Miguel Quartin

## Abstract

We infer the infection fatality ratio (IFR) of SARS-CoV-2 in Brazil by combining three datasets. We compute the prevalence via the population-based seroprevalence survey EPICOVID19-BR. For the fatalities we obtain the absolute number using the public Painel Coronavírus dataset and the age-relative number using the public SIVEP-Gripe dataset. The time delay between the development of antibodies and subsequent fatality is estimated via the SIVEP-Gripe dataset. We obtain the IFR via Bayesian inference for each survey stage and 27 federal states. We include the effect of fading IgG antibody levels by marginalizing over the time after contagion at which the test gives a negative result with a flat prior on the interval [40, 80] days. We infer a country-wide average IFR (maximum posterior and 95% CI) of 0.97% (0.82–1.14%) and age-specific IFR: 0.028% (0.024–0.036%) [< 30 years], 0.21% (0.17–0.25%) [30–49 years], 1.06% (0.88–1.31%) [50–69 years], 2.9% (2.5–3.7%) [≥ 70 years].

## INTRODUCTION

The infection fatality ratio (IFR) – the ratio between the number of deaths from a disease and the number of infected individuals (irrespective of developing symptoms) – is one of the most important quantities of any new disease. An accurate estimate of the IFR is usually a challenge before the end of a pandemic, being subject to many possible sources of biases.^1,2^ Nevertheless, the IFR has direct implications on the amount of resources and effort that should be allocated to prevent the spread of the disease and on steer policy-making in general. For instance using the United States as reference, Perlroth et al.^3^ concluded that an IFR below 1% makes school-closures and social distancing not cost-effective.

In order to estimate the IFR one needs not only an estimate of the number of deaths, but also of the total infected population, and then to compare both within the same time period. It is, therefore, a difficult task as many cases are asymptomatic or develop only mild symptoms and are often unaccounted for. It is also hampered due to the lack of testing in many countries.^4^

The total number of deaths during an epidemic can be biased by the mislabeling of undiagnosed fatalities. To circumvent this possibility, one can rely on statistical estimates from the study of the excess deaths in a given period of time. In the case of COVID-19 this method is being pursued by many groups,^5–7^ including the mainstream media,^8–10^ as a method which is complementary to the officially reported numbers. However, this approach invariably suffers from important modeling uncertainties.^6^ This may be especially true during the current pandemic which has seen an unprecedented amount of disruption of economic activity and social behavior, which includes a large fraction of the population undertaking social distancing measures.^11^

One of the first detailed analyses of the IFR of COVID-19 was based on around 70 thousand clinically diagnosed cases in China. After adjusting for demography and under-ascertainment Verity et al. arrived at the estimate of 1.38% (95% CI: 1.23–1.53%).^12^ In France, a study recently modeled both death and hospital data and estimated the IFR to be 0.5% (95% CI: 0.3–0.9%).^13^ Another model-based investigation arrived at an IFR of 0.8% (95% CI: 0.45–1.25%).^14^ A meta-analysis of 25 IFR studies found an IFR of 0.68% (95% CI: 0.53–0.82%).^15^ A compartmental model for IFR in China and Europe using surveillance data resulted in the following estimates: Hubei, China 2.9% (95% CI: 2.4%–3.5%); Switzerland 0.5% (95% CI: 0.4%–0.6%); Lombardy, Italy 1.4% (95% CI: 1.1%–1.6%).^16^ Finally, relying on antibody screening of Danish blood donors – all younger than 70 and healthy – the IFR was estimated to be less than 0.21% at 95% CL.^17^ These results hint at a possible large variation in IFR values around the globe, although data from different countries were reported to be highly heterogeneous.

In Brazil, the focus of this work, the IFR was recently estimated with models. Results varied substantially between two different groups. A Brazilian team found that it should be much lower than the first estimates, around 0.3%.^18^ On the other hand, a report by the group at Imperial College London estimated much higher values for the 16 Brazilian states they considered,^19^ which, combined, suggest an overall IFR of 0.9%.

The incompatible estimates above highlight the need for a careful consideration of the biases in the IFR estimates for COVID-19. A possible solution to mitigate biases is to base the estimation of the IFR on a large representative random serology study in the population. One such study – conducted in Geneva, Switzerland, with 2766 participants – found that for every reported COVID-19 case there were another 10.6 unreported ones,^20^ a large discrepancy which again stresses the difficulties that models have to deal with. The same group reported an IFR of 0.64% (95% CI: 0.38–0.98%).^21^ A much larger survey with 61075 participants was conducted in Spain, but IFR estimates were not reported.^22^

A large random seroprevalence study was performed in Brazil by the EPICOVID19-BR team^23–25^ which aimed to test 250 individuals in each of the 133 selected large sentinel cities. It was an extension to the whole country of a smaller regional set of surveys in the Rio Grande do Sul state^26^ and has so far been carried out in 4 stages using the Wondfo lateral flow test for immunoglobulin M and G antibodies against SARS-CoV-2. Here, we consider data relative to the first three stages. The first stage was conducted between May 14 and 21, 2020, but did not reach its target number of samples, and in only 90 of the 133 cities at least 200 tests were performed. The total number of tests in all cities was 25025. Round 2 was conducted from June 4 to 7 and reached over 200 tests in 120 cities. Considering all cities a total of 31165 individuals were tested. Round 3 was performed between June 21 and 24 and made over 200 tests in all 133 cities for a total of 33207 tests. The total number of tests in all rounds was 89397.

The COVID-19 pandemic has strongly affected Brazil.^27^ The federal government response has been heavily criticized,^28^ and in December 2020 the number of confirmed cases and deaths crossed 7 million and 180 thousand, respectively, second only to the USA in the raw number of deaths. Furthermore, strong ethnic end regional variations in hospital mortality were found, casting doubts on the availability of public health care for the sections of society that cannot afford private care.^29^ This daring situation motivates even further the need for an estimation of the IFR which is as accurate as possible in order to trigger an adequate political response to the crisis.

## RESULTS AND DISCUSSION

Figure 1 summarizes how the three datasets used in this work where combined in order to estimate the IFR. Our estimations of the IFR for Brazil are given in Table I and Figure 2, while the ones for the individual states (combining all rounds) in Figure 3. We note significant statistical tension in the data of Roraima (RR) (details in the Supplementary Materials). We, therefore, consider its IFR estimate unreliable, but due to its small population it has an insignificant impact on the IFR estimates at the country-level. The numerical results for all the states and for the three rounds separately can be found in the Supplementary Materials. The confidence intervals are computed by combining the statistical sources of error and including the non-Gaussian nature of the distributions. Since we marginalize the posterior on the IFR over the IgG fading time *T*, our estimation is robust against the uncertainty on *T* and the confidence interval includes the effect of the correlation between IFR and *T* (see Materials and Methods).

**Table I.** Cumulative IFR for Brazil (maximum of probability distribution and 95%CI).

| Brazil | date | antibody prevalence (%) | IFR (%) |
| --- | --- | --- | --- |
| Round 1 | 17 May 2020 | 2.62(2.04–3.23) | 0.96(0.77–1.26) |
| Round 2 | 5 Jun 2020 | 3.79(3.22–4.40) | 0.97(0.75–1.17) |
| Round 3 | 22 Jun 2020 | 3.81(3.27–4.39) | 1.16(0.76–1.37) |
| All ages | all rounds | – | 0.97(0.82–1.14) |
| < 30 years | all rounds | – | 0.028(0.024–0.036) |
| 30–49 years | all rounds | – | 0.21(0.17–0.25) |
| 50–69 years | all rounds | – | 1.06(0.88–1.31) |
| ≥ 70 years | all rounds | – | 2.9(2.5–3.7) |

**Figure 1.**
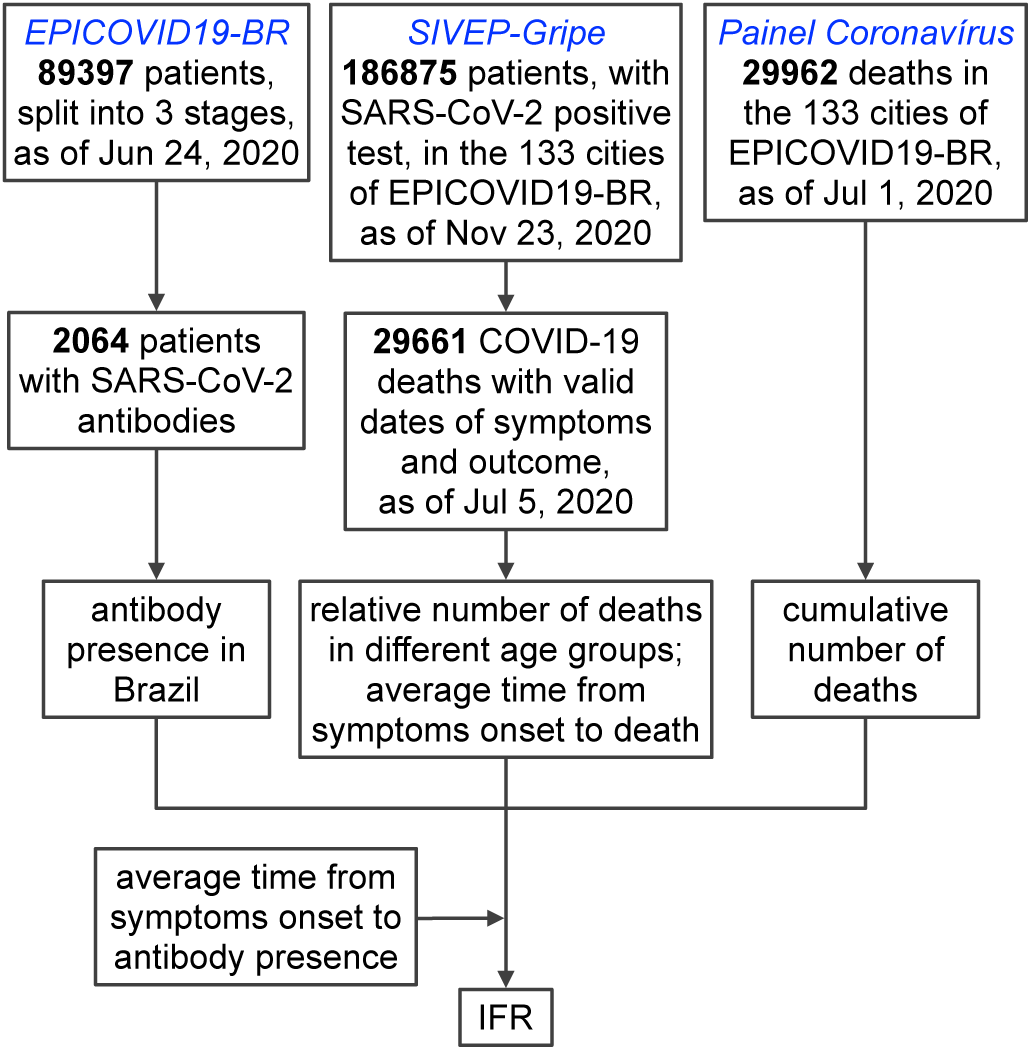
Flowchart of data used in this study.

**Figure 2.**
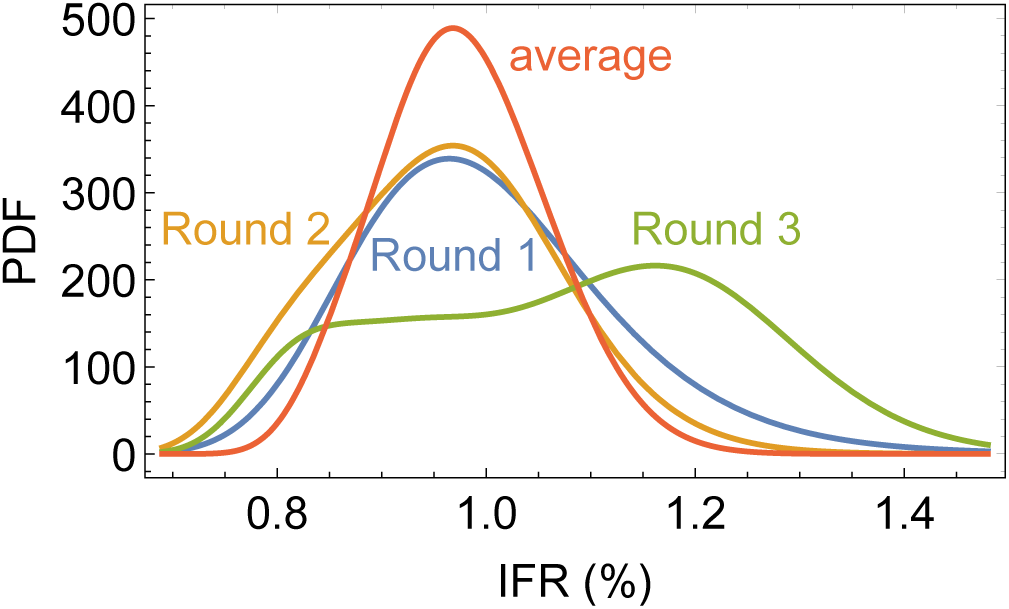
IFR posterior PDF for Brazil for each of the 3 rounds and all rounds combined.

**Figure 3.**
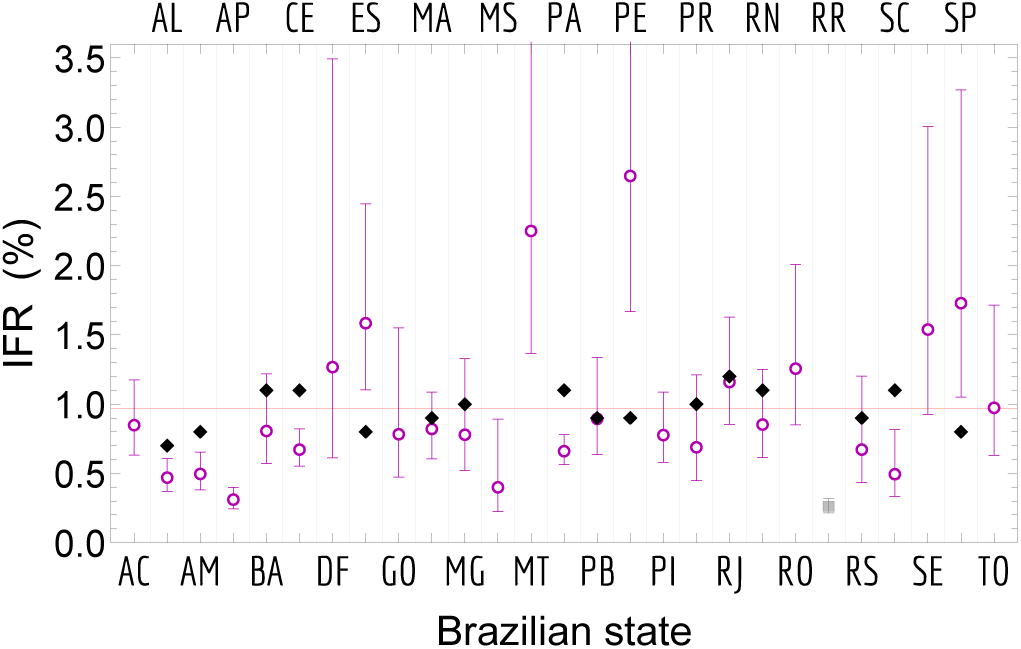
Combined IFR using all 3 rounds. Shown is the maximum posterior and 95%CI. The black dots represent model-based results by the Imperial College COVID-19 Response Team.19 The horizontal red line is the IFR estimate for Brazil given in Table I.

Our overall estimate of the IFR of 0.97% (95% CI: 0.82–1.14%) is in agreement with some, but not all, of the previous world estimates discussed earlier. In particular, at the country level, our combined estimate agrees with the one by the Imperial College COVID-19 Response Team,^19^ even though at the state level we find several disagreements between their values and our 95% CIs, see Figure 3.

It is well known that the IFR of COVID-19 strongly depends on age. As the Painel Coronavírus dataset does not provide age information, we adopt the Painel Coronavírus dataset for the absolute number of fatalities and the much richer but hospital-specific SIVEP-Gripe dataset for the relative number of fatalities for the various age bins. We henceforth split the total population into 4 age groups (in years): *<*30, 30–49, 50–69 and *≥* 70. We find that not only the IFR but also the prevalence increases with age. For Round 3, for instance, we find *p*_*a*_ = 3.7%, 4.9%, 6.4%, 8.4% for the age groups *<*30, 30–v 49, 50–69 and*≥* 70, respectively. For Round 1 and 2 the pattern is similar. The result for the IFR is summarized in Table I. The trend of a rapid increase of the IFR with age agrees well with previous findings^30,31^ and urges authorities to promote measures to protect elderly people from SARS-CoV-2 exposure.

In Figure 4, instead of marginalizing over *T*, we show the Brazilian IFR as a function of *T*. For *T* > 80 days the results converge and the IFR remains unchanged. We also find that the effect of neglecting the time delay *τ*_*ad*_ between the development of antibodies and subsequent fatality results in an underestimate of the IFR of around 0.14%, roughly a 1.7 standard-deviation shift. For instance, the country-wide IFR for all ages changes from 0.97% (95% CI: 0.82–1.14%) to 0.83% (95% CI: 0.74– 0.95%).

**Figure 4.**
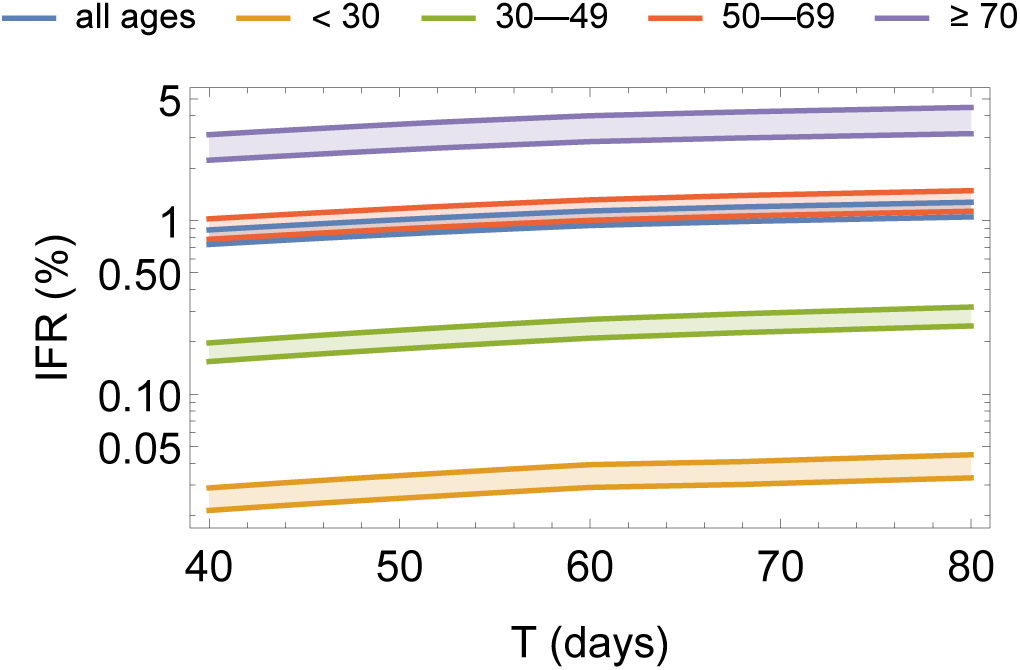
IFR for Brazil for all rounds combined as a function of IgG fading time *T* **and age**. Shown is the 95% CI using the logarithmic scale. For *T >* 80 the results converge and the IFR remains unchanged.

Our estimate features a small 8% standard deviation including the uncertainty on the IgG fading time *T*, but it may suffer from the following systematic biases. First, not all COVID-19 related deaths may be registered in Painel Coronavírus. One expects this to happen for out-of-hospital fatalities and be stronger in the poorest areas with a less present health care infrastructure. Some reports indeed claim a non-negligible number of undiagnosed respiratory deaths in 2020.^32^ However, as we are analyzing the 133 large sentinel cities that entered the EPICOVID19-BR survey this bias is not expected to be significant. Its effect is, nonetheless, the underestimation of the IFR.

Regarding the computation of the IFR as a function of age, a potential bias could arise from the fact that the age distribution of the SIVEP-Gripe dataset may not be representative of the one of the overall population as some age groups may have a higher tendency to present themselves at the hospital and enter the SIVEP-Gripe dataset.

Another potential bias comes from the fact that the time in Painel Coronavírus is not the actual time of death but rather the time of notification. In order to alleviate this issue and also average out oscillations due to week-ends, we smooth the d*n*_*d*_*/*d*t* data according to a forward 20-day moving average which corresponds to shifting on average the time of deaths by 10 days earlier. This figure is justified by a direct comparison between Painel Coronavírus and SIVEP-Gripe and by estimates of the delay between time of death and notification^33,34^ (details in the Supplementary Materials).

Next, the SIVEP-Gripe dataset is biased towards cases with severe symptoms. Indeed, there is a significant number of cases that are hospitalized when symptoms are notified (see Supplementary Materials). We took this into account via a delay parameter 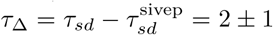 days (see Table II) which models the time that a patient takes to go from symptoms onset to severe symptoms (details in the Supplementary Materials). Had we set *τ*_Δ_ = 0, we would have obtained a relative 2% lower estimate.

**Table II.**
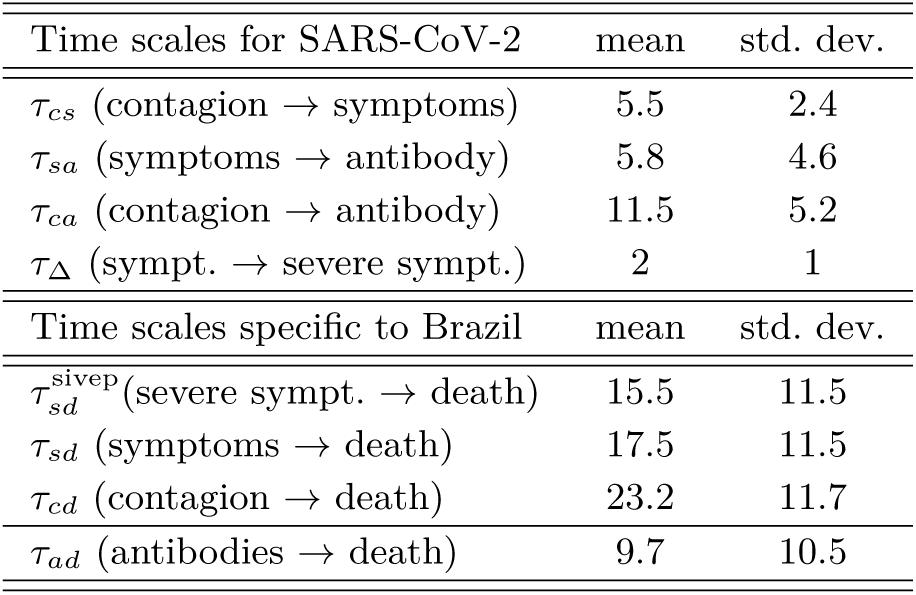
Time scales used in the analysis (in days).

| Time scales for SARS-CoV-2 | mean | std. dev. |
| --- | --- | --- |
| $\tau_{cs}$ (contagion → symptoms) | 5.5 | 2.4 |
| $\tau_{sa}$ (symptoms → antibody) | 5.8 | 4.6 |
| $\tau_{ca}$ (contagion → antibody) | 11.5 | 5.2 |
| $\tau_{\Delta}$ (sympt. → severe sympt.) | 2 | 1 |
| Time scales specific to Brazil | mean | std. dev. |
| $\tau_{sd}^{\text{sivep}}$ (severe sympt. → death) | 15.5 | 11.5 |
| $\tau_{sd}$ (symptoms → death) | 17.5 | 11.5 |
| $\tau_{cd}$ (contagion → death) | 23.2 | 11.7 |
| $\tau_{ad}$ (antibodies → death) | 9.7 | 10.5 |

Finally, the participants of the study may not be fully representative of the whole population of Brazil. The EPICOVID19-BR survey took several measures to mitigate enrollment bias, as discussed by Hallal et al.^25^ For instance, for each city 25 census tracts were selected and in each one 10 households were selected at random. In each one a resident was also selected at random from a listing of members. If the selected individual refused participation another resident was selected at random and if they also refused (or if the residents were absent) the team moved into the neighboring household on the right. As discussed in the Supplementary Materials the age and ethnic distribution of the participants follow well the distributions in Brazil, with a small undersampling of under 20s and people of white ethnicity. Nevertheless since the survey considered only 133 large sentinel cities the overall IFR we computed is relative to these cities, which amount to 35.5% of the Brazilian population. One may nevertheless speculate that the IFR could be different in smaller cities and rural or poorer areas.

The IFR of COVID-19 depends not only on the patient’s age, as discussed earlier, but also on their health.^35^ Our IFR estimate should, therefore, be contextualized to the Brazilian population. To this end, a reasonable proxy for the overall health of a country is life expectancy, and the lower socioeconomic development of Brazil is reflected into a lower life expectancy as compared to, for example, Europe—76.0 years as compared with 80.9 years, as of 2017.^36,37^

As new medications and treatment protocols for the disease are discovered and become available it is hoped that the IFR will decrease. Since our data comes from the first months of the pandemic, our results therefore also set a baseline for future comparisons of the fight against COVID-19 in Brazil.

Concluding, we hope that our careful evaluation of the IFR in Brazil will help reinforce, at the federal, state and municipal levels, the seriousness of the COVID-19 pandemic and the urgency of taking the proper actions in order to reduce its societal and economic impact.

## MATERIALS AND METHODS

As summarized by Figure 1, in order to estimate the IFR we make use of three complementary datasets. We compute the percentage *p*_*a*_(*t*) of Brazilians that have been infected by SARS-CoV-2 at the city, state and Brazilian levels via the EPICOVID19-BR data. We robustly correct for false positive and negative rates^39^ and combine prevalences from different cities without neglecting the non-Gaussian nature of the distributions (details in the Supplementary Materials). The result is shown in Figure 5 and in Table I (the full numerical tables can be found in the Supplementary Materials). We note a sharp increase in prevalence between rounds 1 and 2, and a subsequent stabilization between rounds 2 and 3. The state of Pará (PA) exhibits a sharp decrease in prevalence in round 3.

**Figure 5.**
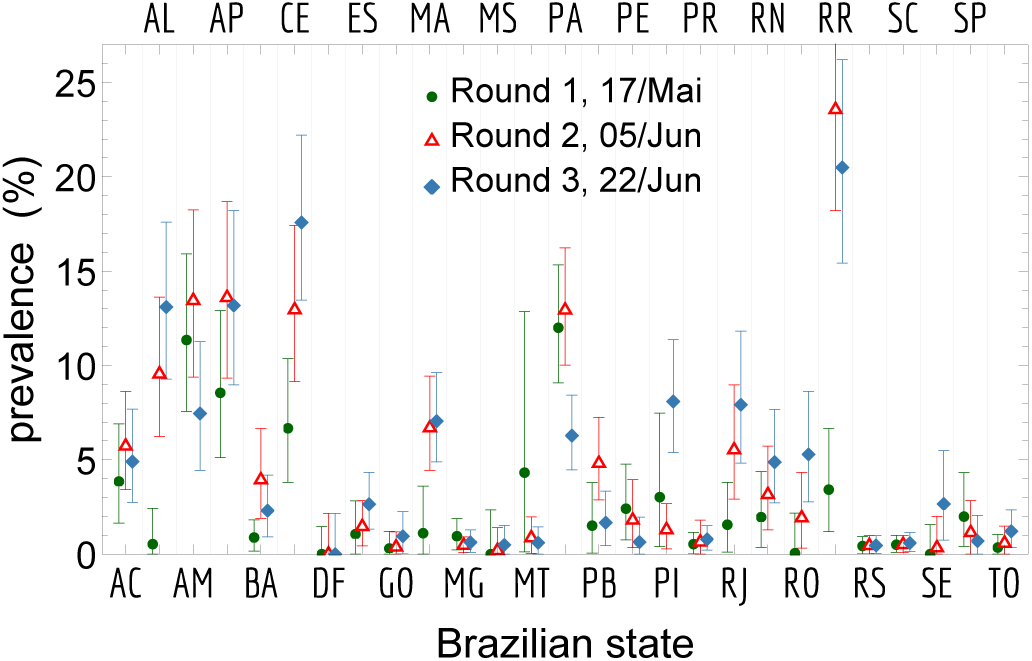
Prevalence of COVID-19 antibodies in each of the 27 Brazilian states in the 3 rounds of the EPICOVID19-BR survey. Shown is the maximum posterior and 95% CI. The state acronyms are AC: Acre, AL: Alagoas, AM: Amazonas, AP: Amapá, BA: Bahia, CE: Ceará, DF: Distrito Federal, ES: Espírito Santo, GO: Goiás, MA: Maranhão, MG: Minas Gerais, MS: Mato Grosso do Sul, MT: Mato Grosso, PA: Pará, PB: Paraíba, PE: Pernambuco, PI: Piauí, PR: Paraná, RJ: Rio de Janeiro, RN: Rio Grande do Norte, RO: Rondônia, RR: Roraima, RS: Rio Grande do Sul, SC: Santa Catarina, SE: Sergipe, SP: São Paulo, TO: To-cantins.

We obtain the absolute number of fatalities via the public Painel Coronavírus dataset.^40^ Painel Coronavírus is the Brazilian reference to keep track of the pandemic at the federal level and provides the deaths by COVID-19 with their geographic location. Reports from Long *et al*.^41^ and more recently by Grzelak et al.^42^ show that IgG levels fade in recovered patients on a timescale of a few months, which was also suggested by the results relative to the first 2 rounds of EPICOVID19-BR.^25^ Moreover, preliminary results from the recent fourth round of EPICOVID19-BR exhibit a large decrease in seroprevalence in the country,^43^ which is consistent with a short window of detectability. For this reason we consider here a detectability window *T* and thus the number of fatalities relative only to such a window, which is equivalent to assume a sharp drop of IgG levels after *T* days. In order to take into account the uncertainty on *T*, we marginalize our results with respect to this parameter adopting a broad flat prior on the interval [40, 80] days. The upper extremum of 80 days is justified by comparing the seroprevalence of the fourth and third rounds of EPICOVID19-BR (details in the Supplementary Materials). The lower limit was chosen in a conservative way as half of the upper limit.

We cannot compute the IFR directly via the ratio of *p*_*d*_ and *p*_*a*_ because, at a given time 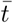, there are patients that developed antibodies but did not die yet from the disease.^44^ In order to estimate the time delay *τ*_*ad*_ between the development of antibodies and subsequent fatality we use the public SIVEP-Gripe dataset (“Sistema de Informação da Vigilância Epidemiológica da Gripe”), a prospectively collected respiratory infection registry data that is maintained by the Ministry of Health for the purposes of recording cases of Severe Acute Respiratory Syndrome (SARS) across both public and private hospitals. The SIVEP-Gripe dataset contains the dates of symptoms onset and death for patients with SARS-CoV-2 positive RT-PCR test, together with their geographic location and age, which allow us to estimate the time delay *τ*_*sd*_ between the development of symptoms and subsequent fatality. We also make use of an empirical distribution between the first symptoms and the development of antibodies^45^ to estimate the mean time-delay *τ*_*sa*_ between both events. Together, these estimates allow us to obtain the time-delay *τ*_*ad*_ ≃*τ*_*sd*_ −*τ*_*sa*_. For the whole Brazil we find *τ*_*ad*_ ≃9.7 days. Table II summarize all the estimated time-delays which are used in our calculations (details in the Supplementary Materials).

Using this combined information we can then finally compute the IFR at the state and country levels

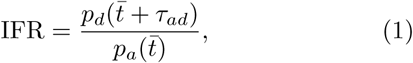

where 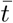 is the time of a given EPICOVID19-BR phase.

## Data Availability

The data and codes used for this work are made publicly available.

https://github.com/mquartin/covid19-ifr-br

## Data availability

All data needed to evaluate the conclusions in the paper are present in the paper and/or the Supplementary Materials.

## Code availability

The Wolfram Mathematica codes and the data used for this work are available at: www.github.com/mquartin/covid19-ifr-br.

## Acknowledgements

VM’s research is partially supported by the Brazilian research agencies CNPq and FAPES. MQ’s research is partially supported by the Brazilian research agencies CNPq and FAPERJ. This project has not received any funding.

## Author contributions

VM and MQ both contributed equally to this work.

## Competing interests

The authors declare no competing interests.

## Supplementary Materials

Materials and Methods

Sections S1 to S7

Figs. S1 to S9

Table S1

## Notes

### Competing Interest Statement

The authors have declared no competing interest.

### Funding Statement

No funding was provided.

### Summary of Updates

v2: updated to include the effect of fading IgG levels. v3: updated to include the IFR for various age bins.

