## Supplementary materials for "A Bayesian estimate of the COVID-19 infection fatality ratio in Brazil based on a random seroprevalence survey"

### CONTENTS

Materials and Methods 1

S1. Antibody prevalence 1

S2. Infection fatality ratio 2

S3. Uncertainty estimation 4

S4. IFR for the three EPICOVID19-BR rounds 5

S5. IFR as a function of age 5

S6. Representativeness of the survey 5

S7. Full numerical results 6

References 8

### Materials and Methods

#### S1. ANTIBODY PREVALENCE

The antibody presence in the population $p_a$ was directly assessed via the EPICOVID19-BR serological survey, which adopted the One Step COVID-19 Test from Wondfo. The test was subjected to analysis of its specificity and sensitivity by different groups including the manufacturer. Specificity and sensitivity are in turn related directly to the false positive rate (FPR) and false negative rate (FNR), respectively. The EPICOVID19-BR team consolidated different analyses of this test and arrived at a FPR = 1.0% (95% CI: 0.3–2.2%) and FNR = 15.2% (95% CI: 12.2–18.6%).[26]

If both FPR and FNR were zero, each test outcome would be drawn from a simple Binomial distribution with probability $p_a$. Therefore $p_a$ would be distributed according to the conjugate-prior distribution, which for $n_{\rm tot}$ tests with $n_{\rm yes}$ positive detections, would be a Beta distribution $\mathcal{B}[\alpha,\beta](p_a)$ with $\alpha = n_{\rm yes}+1$ and $\beta = n_{\rm tot} - n_{\rm yes} + 1$. A non-zero FPR and FNR can be taken into account with a simple change of variables $p_a^{\rm obs} \to p_a^{\rm true}(1-{\rm FNR}-{\rm FPR})+{\rm FPR}$. The distribution for $p_a^{\rm true}$ can then be computed numerically to arbitrary precision, but it can also be approximated to great accuracy by a Pearson type I distribution, sometimes referred to as the 4-parameter Beta distribution, which, in our case, can be simply thought of as a displaced Beta distribution:

$$\mathcal{B}[\alpha,\beta](p_a+p_{\rm min})\,, \tag{1}$$

with $p_{\rm min} = {\rm FPR}/(1-{\rm FNR}-{\rm FPR})$ is the displacement, $\alpha = n_{\rm yes}+1$ and $\beta = n_{\rm tot}(1-{\rm FNR}-{\rm FPR})-{\rm n_{yes}}+1$. Since this mathematically allows $p_a < 0$, it must be multiplied by a uniform (or top-hat) prior enforcing $p_a \geq 0$, to wit $\mathcal{U}[0,1](p_a)$.

In order to combine the prevalence results in different cities and get results for a given state or for the whole country we proceed as follow. The combined prevalence is given by:

$$p_a^{\rm combined} = \frac{N_a}{\rm TotPop} = \frac{\sum_{{\rm city}\ i} p_{a,i}{\rm pop}_i}{\rm TotPop} = \sum_{{\rm city}\ i} p_{a,i}{\rm fpop}_i. \tag{2}$$

In the above, $N_a$ is the number of people with antibodies in the combination of $N$ cities, TotPop is the total combined population, $p_{a,i}$ and ${\rm pop_i}$ is the prevalence and population in city $i$, respectively, and ${\rm fpop}_i$ is the fraction of the total population considered in city $i$. This means that the distribution for $p_a^{\rm combined}$ is technically computed as a transformation of many variables $p_{a,1},\ldots,p_{a,N}$ into one variable $p_a^{\rm combined}$. This leads to the following challenging $N$-dimensional integral for the state $s$:

$$\mathcal{L}(p_{a,s}) = \mathcal{N}\int {\rm d}^N p_{a,j}\,\delta\Big[p_{a,s} - \sum_{{\rm city}\ j} p_{a,j}{\rm fpop}_j\Big]\prod_{{\rm city}\ i}\mathcal{L}_i(p_{a,i}), \tag{3}$$

where $\delta$ is the Dirac delta function and $\mathcal{N}$ a normalization constant. We have computed such integral for all states in which $N \leq 4$ and found out that this exact integral can be accurately approximated once again by a displaced Beta distribution.

The displacement is exactly as above since it comes, after all, from the FPR and FNR, which is assumed to be the same in all cities as the same test was employed. In order to find the values of $\alpha$ and $\beta$ we require that the Beta distribution has the correct mean $\mu$ and variance $V$. For a standard Beta distribution these first 2 moments

* Contributed equally

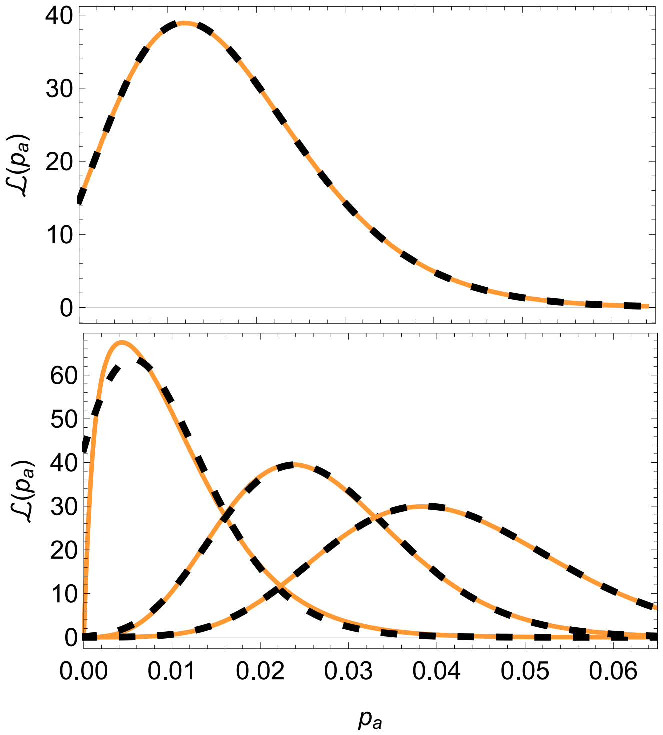


Figure S1. **Comparison of the Beta distribution approximations (dashed line in black) and exact results (solid line in orange).** *Top*: using Eq. (1) for a single city, assuming 5 positive results out of 250 tests and the FNR and FPR discussed in the text. There is no discernible difference. *Bottom*: using Eqs. (4) and (5) for the states AC, AL and PE in Round 1. There is only a small difference in the cases where the maximum likelihood is very small ($p_a < 0.015$), otherwise the approximation is indistinguishable.

of the distribution define both parameters univocally:

$$\alpha = \frac{\mu}{V}\big(-V+\mu-\mu^2\big)\,, \quad \beta = \frac{1-\mu}{V}\big(-V+\mu-\mu^2\big)\,. \quad (4)$$

Since our approximation consists on a truncated displaced Beta – due to the prior $\mathcal{U}[0,1](p_a^{\text{combined}})$ – the above relation is only approximately valid, and it gets worse in the cases with few positive results because the prior becomes informative. We therefore use it only as a first guess and find $\alpha$ and $\beta$ numerically demanding that the final $\mu$ and $V$ have the correct values. The combined $\mu$ and $V$ in turn are easily obtained using Eqs. (2) and (4) and the properties of the mean and variance of a sum of independent random variables. To wit, if $\mu_i$ and $V_i$ are the mean and variance of city $i$, we get:

$$\mu = \sum_{\text{city } i} \mu_i \, \text{fpop}_i\,, \quad V = \sum_{\text{city } i} V_i \, \text{fpop}_i^2\,. \quad (5)$$

The resulting approximation is very good, and in particular it leads to medians (95% confidence interval widths) which differ on average from the exact result by a relative amount of just $\simeq 3\%$ (0.5%). Moreover, there are no severe outlier cases for this approximation, and for the whole range of states and rounds the largest discrepancy of medians was 6%. Figure S1 shows the excellent performance of the Beta distribution approximation.

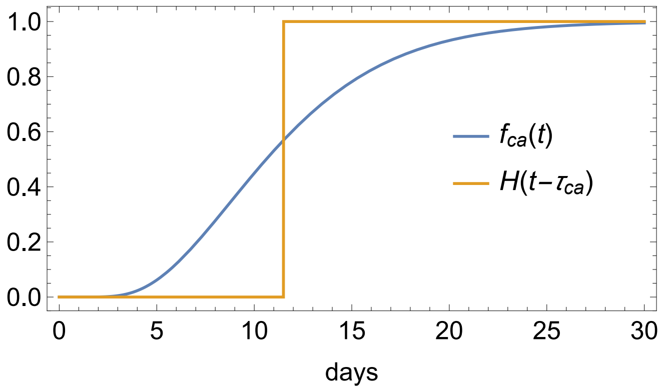


Figure S2. **Cumulative probability distribution of IgG and/or IgM presence as a function of days after contagion.**

### S2. INFECTION FATALITY RATIO

In order to estimate the IFR we need to relate the cumulative number $n_+(t)$ of COVID-19 cases to the cumulative number $n_a(t)$ of patients with SARS-CoV-2 antibodies and the cumulative number $n_d(t)$ of non-survivors. To this end we need to estimate the average time $\tau_{ca}$ from contagion to antibody presence and the average time $\tau_{cd}$ from contagion to death, and then compute the time-delay between antibodies and death $\tau_{ad} \simeq \tau_{sd} - \tau_{sa} \simeq \tau_{cd} - \tau_{ca}$.

We obtain $\tau_{ca}$ from the mean of the convolution $\mathrm{d}f_{ca}/\mathrm{d}t$ of the distributions of the time from contagion to symptoms onset $\mathrm{d}f_{cs}/\mathrm{d}t$ and from symptoms onset to antiviral immunoglobulin G (IgG) and/or M (IgM) presence $\mathrm{d}f_{sa}/\mathrm{d}t$:

$$\frac{\mathrm{d}f_{ca}(t)}{\mathrm{d}t} = \int_0^t \mathrm{d}\bar{t}\, \frac{\mathrm{d}f_{sa}(\bar{t})}{\mathrm{d}\bar{t}}\, \frac{\mathrm{d}f_{cs}}{\mathrm{d}t}(t-\bar{t})\,, \quad (6)$$

$$f_{ca}(t) = \int_0^t \mathrm{d}\bar{t}\, \frac{\mathrm{d}f_{ca}(\bar{t})}{\mathrm{d}\bar{t}} \simeq H(t-\tau_{ca})\,, \quad (7)$$

where in the last equation we used the instantaneous approximation according to which $\mathrm{d}f_{ca}/\mathrm{d}t \simeq \delta(t-\tau_{ca})$ is a Dirac delta function so that $f_{ca}$ is modeled as a Heaviside step function. The distribution $\mathrm{d}f_{sa}/\mathrm{d}t$ is modeled empirically,[45] while $\mathrm{d}f_{cs}/\mathrm{d}t$ according to a lognormal template.[46] The means $\tau_{ca}$, $\tau_{cs}$ and $\tau_{sa}$ relative to $\mathrm{d}f_{ca}/\mathrm{d}t$, $\mathrm{d}f_{cs}/\mathrm{d}t$ and $\mathrm{d}f_{sa}/\mathrm{d}t$ are reported in Table II of the main text. Note that $\tau_{ca} \simeq \tau_{cs} + \tau_{sa}$. The functions $f_{ca}(t)$ and $H(t-\tau_{ca})$ are shown in Figure S2. $\tau_{sd}$ is in agreement with the average time from symptoms onset to death relative to other countries.[47,48]

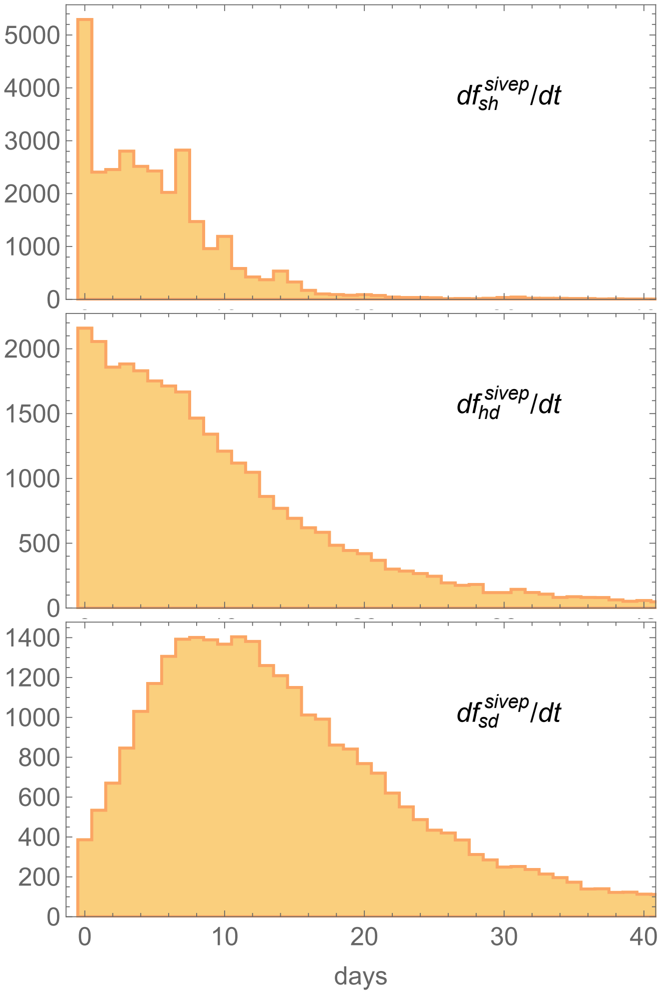


Figure S3. **Distribution of times between different events.** *Top*: days from symptoms onset to hospitalization. *Middle*: days from hospitalization to death. *Bottom*: days from symptoms onset to death.

Similarly, we obtain $\tau_{cd}$ from the mean of the convolution $\mathrm{d}f_{cd}/\mathrm{d}t$ of the distributions of the time from contagion to symptoms onset $\mathrm{d}f_{cs}/\mathrm{d}t$ and from symptoms onset to death $\mathrm{d}f_{sd}/\mathrm{d}t$:

$$\frac{\mathrm{d}f_{cd}(t)}{\mathrm{d}t} = \int_0^t \mathrm{d}\bar{t}\, \frac{\mathrm{d}f_{sd}(\bar{t})}{\mathrm{d}\bar{t}} \frac{\mathrm{d}f_{cs}}{\mathrm{d}t}(t-\bar{t})\,, \tag{8}$$

$$f_{cd}(t) = \int_0^t \mathrm{d}\bar{t}\, \frac{\mathrm{d}f_{cd}(\bar{t})}{\mathrm{d}\bar{t}} \simeq H(t-\tau_{cd})\,. \tag{9}$$

The distribution $\mathrm{d}f_{sd}/\mathrm{d}t$ is modeled empirically using the data from the SIVEP-Gripe dataset at the Brazilian and state levels as described below. Note that $\tau_{cd} \simeq \tau_{cs} + \tau_{sd}$.

The data from SIVEP-Gripe is biased towards cases with severe symptoms. Indeed, there is a significant number of cases that are hospitalized when symptoms are notified (see Figure S3, top panel). In order to take this into account we introduce a delay parameter $\tau_\Delta$ (see Table II of the main text) which models the time that a patient

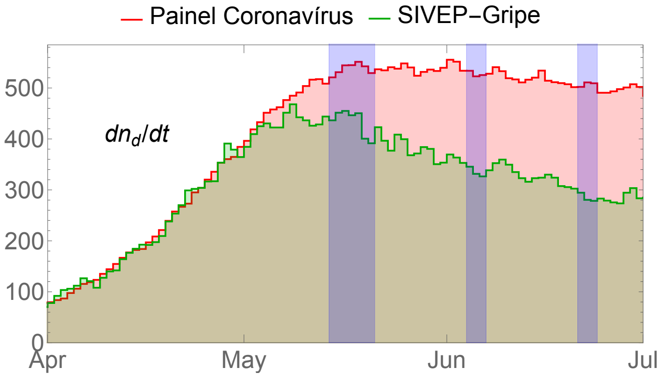


Figure S4. **New deaths by COVID-19 for the 133 cities that entered the EPICOVID19-BR survey.** The purple windows mark the three survey rounds. The data from Painel Coronavírus was smoothed according to a forward 20-day moving average which corresponds to shifting on average the time of deaths by 10 days earlier.

takes on average to go from symptoms onset to severe symptoms:

$$\frac{\mathrm{d}f_{sd}}{\mathrm{d}t}(t) = \int_0^t \mathrm{d}\bar{t}\, \frac{\mathrm{d}f_{sd}^{\text{sivep}}(\bar{t})}{\mathrm{d}\bar{t}} \frac{\mathrm{d}f_\Delta}{\mathrm{d}t}(t-\bar{t}) \simeq \frac{\mathrm{d}f_{sd}^{\text{sivep}}}{\mathrm{d}t}(t-\tau_\Delta), \tag{10}$$

where we again used the instantaneous approximation for the distribution $\mathrm{d}f_\Delta/\mathrm{d}t$ and $\tau_\Delta$ is its mean. We estimate $\tau_\Delta = 2$ days.[49] This means that $\tau_{sd} = \tau_{sd}^{\text{sivep}} + \tau_\Delta$. The distribution $\mathrm{d}f_{sd}^{\text{sivep}}/\mathrm{d}t$ at the Brazilian level is shown in Figure S3 (bottom panel). In the computation of the IFR we will adopt the median of $\mathrm{d}f_{sd}^{\text{sivep}}/\mathrm{d}t$ as a robust measure of $\tau_{sd}^{\text{sivep}}$.

We can now relate the cumulative number $n_+(t)$ of COVID-19 cases to the cumulative number $n_a(t)$ of patients with SARS-CoV-2 antibodies and the cumulative number $n_d(t)$ of non-survivors:

$$n_a(t) = \int_{t_0}^t \mathrm{d}\bar{t}\, \frac{\mathrm{d}n_+(\bar{t})}{\mathrm{d}\bar{t}} f_{ca}(t-\bar{t}) \simeq n_+(t-\tau_{ca})\,, \tag{11}$$

$$n_d(t) = \mathrm{IFR} \int_{t_0}^t \mathrm{d}\bar{t}\, \frac{\mathrm{d}n_+(\bar{t})}{\mathrm{d}\bar{t}} f_{cd}(t-\bar{t}) \simeq \mathrm{IFR}\, n_+(t-\tau_{cd}), \tag{12}$$

where we assumed that IFR is constant. The number of fatalities is obtained via the public Painel Coronavírus dataset. Figure S4 shows the new deaths $\mathrm{d}n_d/\mathrm{d}t$ for the 133 cities that entered the EPICOVID19-BR survey. From $\mathrm{d}n_d/\mathrm{d}t$ we can compute the cumulative number of deaths $n_d(t)$.

We can now use the previous two equations to compute the IFR:

$$\mathrm{IFR} = \frac{n_d(t)}{n_+(t-\tau_{cd})} = \frac{n_d(\bar{t}+\tau_{cd}-\tau_{ca})}{n_a(\bar{t})} \simeq \frac{n_d(\bar{t}+\tau_{sd}-\tau_{sa})}{n_a(\bar{t})} = \frac{p_d(\bar{t}+\tau_{ad})}{p_a(\bar{t})}, \tag{13}$$

where $\bar{t}$ is the time relative to the measurement of $n_a$ and in the last equation we divided numerator and denominator by the number $n_{\rm pop}$ of inhabitants according to the 2019 official population values,[50] $p_d = n_d/n_{\rm pop}$. As explained earlier, we estimate $p_a$ using EPICOVID19-BR data and $p_d$ using the Painel Coronavírus data shown in Figure S4. We smooth the Painel Coronavírus data according to a forward 20-day moving average that assigns to the time $t_0$ the average value of deaths in the interval $[t_0, t_0{+}19 \text{ days}]$, which corresponds to shifting on average the time of deaths by 10 days earlier. This figure is justified by a direct comparison between Painel Coronavírus and SIVEP-Gripe and by estimates of the delay between time of death and notification. Indeed, for many reasons, deaths that happen at the time $t_0$ are reported at a later time that we estimate according to a flat distribution in the above mentioned interval.[33,34]

As discussed in the main text, because of fading IgG levels, we consider a detectability window $T$ and thus the number of fatalities relative only to such a window. Specifically, we limit the number of deaths between the delayed time $\bar{t} + \tau_{ad}$ and $T$ days earlier. We treat $T$ as a nuisance parameter which takes values in the interval $[40, 80]$ days.

We estimate the IFR at the state and Brazilian level. This means that in Eq. (13) $n_d$, $\tau_{sd}$ and $n_{\rm pop}$ are calculated accordingly.

### S3. UNCERTAINTY ESTIMATION

Our results are given in terms of the maxima of the probability distributions and highest density intervals. The full probability distribution of $p_a$ was already discussed in Section S1.

Regarding $p_d$, we estimate the error by propagating the uncertainty on $n_d$ due to the uncertainty on $\tau_{sd}$. We estimate the error on $\tau_{sd}$ by adding in quadrature the uncertainty on $\tau_\Delta$ and the uncertainty on $\tau_{sd}^{\rm sivep}$. We estimate the former as $\sigma_\Delta = 1$ day[49] and the latter via bootstrapping from the empirical distribution. Systematic uncertainties are discussed in the main text.

Regarding the IFR, we find that the relative error on $p_d$ is smaller than the one on $p_a$ by a factor between 3.5 to 5 depending on the survey round. We can therefore approximate $p_d$ as a fixed variable in Eq. (13). We then take into account the full distribution of IFR due to the uncertainties in $p_a$. The distribution on the IFR is thus the one of the inverse of a truncated Beta distribution, the PDF of which can be written in closed form as:

$$\frac{p_d\,\Gamma(\alpha+\beta)\left(\frac{p_d}{x}+p_{\rm min}\right)^{\alpha}\left(1-p_{\rm min}-\frac{p_d}{x}\right)^{\beta}}{\Gamma(\alpha)\Gamma(\beta)(p_d+p_{\rm min}x)\big[p_d-(1-p_{\rm min})x\big]\left(I_{p_{\rm min}}[\alpha,\beta]-1\right)} \tag{14}$$

for $x > p_d/(1+p_{\rm min})$ and zero otherwise. Here, $x$ is the free variable, in our case the IFR, $I$ is the regularized incomplete Beta function, and $\Gamma$ is the gamma function.

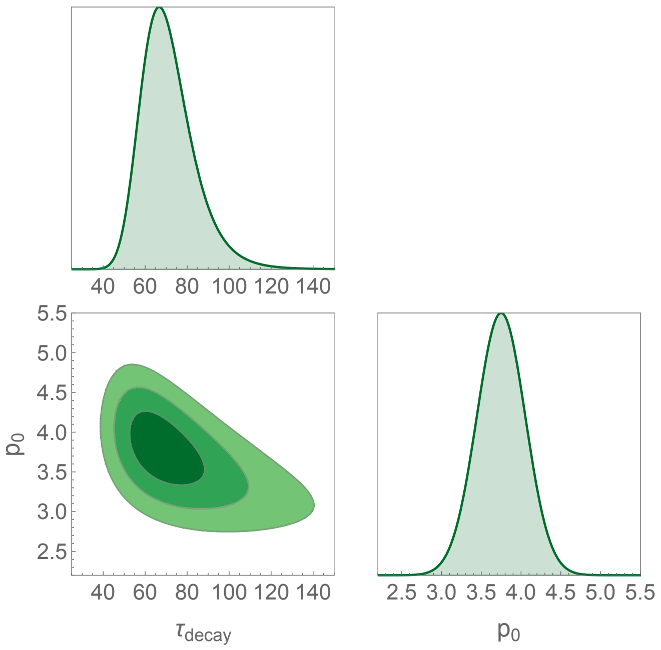


Figure S5. **Full 2D and marginalized 1D posterior distributions on the mean time $\tau_{\rm decay}$ and the normalization $p_0$.** The 2D contours mark the regions with 68.3%, 95.4% and 99.7% of probability.

As discussed earlier and in the main text, because of fading IgG levels, we only consider fatalities between the delayed time $\bar{t} + \tau_{ad}$ and $T$ days earlier. The resulting value of the IFR correlates with $T$ and the fact that $T$ is not precisely known could introduce an important bias in the analysis. In order to robustly overcome this issue we treat $T$ as a nuisance parameter to be integrated over. Specifically, we adopt a broad flat prior $T \in [40, 80]$ days so that the marginalized distribution on the IFR is:

$$\mathcal{P}_{\rm IFR} \propto \int_{40\text{ days}}^{80\text{ days}} \widetilde{\mathcal{P}}_{\rm IFR}\,{\rm d}T\,, \tag{15}$$

where $\widetilde{\mathcal{P}}_{\rm IFR}$ is the distribution of Eq. (14).

In order to justify the upper extremum of 80 days we compare the seroprevalence of the third and fourth rounds of EPICOVID19-BR. Specifically, we can obtain an upper bound on $T$ by assuming an exponential decay with no new infections. We performed a Bayesian analysis for the model $p_a(t) = p_0\, e^{-t/\tau_{\rm decay}}$ where $p_0$, a normalization, is a nuisance parameter and $\tau_{\rm decay}$ gives the mean time after contagion at which the test gives a negative result. The median time of 80 days that we adopted as upper bound corresponds to $80/\ln 2 \simeq 115$ days of mean time, a figure well in agreement with the results shown in Figure S5.

Finally, as the error budget is driven by the uncertainty on $p_a$ and the determinations of $p_a$ during the three rounds are statistically independent, we can combine the rounds by simply multiplying the likelihoods. This combination allows for a more precise estimate of

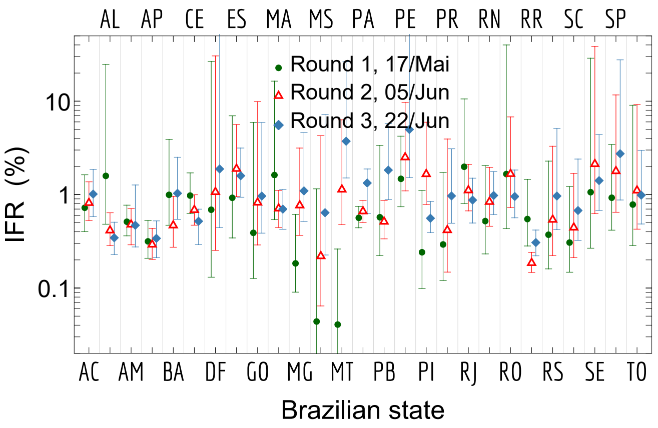


Figure S6. **Separate IFR estimates for each of the 3 rounds for each state (maximum posterior and 95% CI).** The lower statistics leads to larger uncertainties when compared to Figure 3 of the main text.

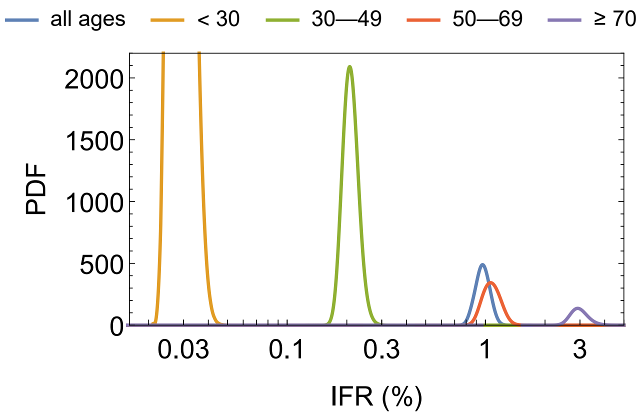


Figure S7. **IFR posterior PDF for Brazil for all rounds combined according to the different age bins considered.** The IFR is in logarithmic scale, as the extreme age groups differ by two orders of magnitude.

the average IFR over all rounds. Clearly, this ignores possible changes of the IFR during the course of the 5 weeks between rounds 1 and 3.

### S4. IFR FOR THE THREE EPICOVID19-BR ROUNDS

Brazil is divided geopolitically into 5 macroregions, the 27 states of which are:

- North: Acre (AC), Amapá (AP), Amazonas (AM), Pará (PA), Rondônia (RO), Roraima (RR), Tocantins (TO);
- Northeast: Alagoas (AL), Bahia (BA), Ceará (CE), Maranhão (MA), Paraíba (PB), Pernambuco (PE), Piauí (PI), Rio Grande do Norte (RN), Sergipe (SE);
- Central-West: Distrito Federal (DF), Goiás (GO), Mato Grosso (MT), Mato Grosso do Sul (MS);
- Southeast: Espírito Santo (ES), Minas Gerais (MG), Rio de Janeiro (RJ), São Paulo (SP);
- South: Paraná (PR), Rio Grande do Sul (RS), Santa Catarina (SC).

Figure S6 shows the IFR estimates for the three EPICOVID19-BR rounds for each state. In most states we note a small increase in IFR on rounds 2 and 3, but specially in the latter. In Roraima (RR) we also note a statistically significant discrepant IFR result in round 2.

### S5. IFR AS A FUNCTION OF AGE

Figure S7 shows the full non-Gaussian distribution of the IFR using (14) and (15) for all rounds obtained combined according to the different age bins considered in this work. The corresponding maxima of the posterior and 95% CI are shown in Table 1 of the main text.

### S6. REPRESENTATIVENESS OF THE SURVEY

Selection bias in serosurveys can introduce important distortions to the inferred results. It is therefore crucial to cross-check the survey data for differential response rates among different subgroups.[51]

Figure S8 depicts the age distributions for the first three rounds of the EPICOVID19-BR survey. We show the relative frequencies of the total number of interviewed people on the survey, of the overall Brazilian population[37] and of positive and negative test results. The distribution of interviewees follow approximately the overall Brazilian distribution with a small bias towards older people, which is likely due to the fact that the survey is based on large satellite cities for which the population pyramid is skewed towards a slightly older population. As can be seen, there is some recruitment bias of roughly 20% against young people under 20s and in favor of over 50s, but otherwise the test distribution follows roughly the overall distribution. We remark that this plot does not take into account the different number of habitants in each city. As discussed in the main text, when taking the city population into account one finds a clear correlation between antibody prevalence and age.

In order to further assess the representativeness of the EPICOVID19-BR survey we show in Figure S9 the ethnic distribution: although people of white (black) ethnicity are slightly under (over) represented, the tested set follows well the national averages, especially considering that EPICOVID19-BR has tested only 133 cities–about 35.5% of the Brazilian population.

Both figures, together with the high response rate of over 50% of the survey, show that EPICOVID19-BR represents well the Brazilian population in terms of age and

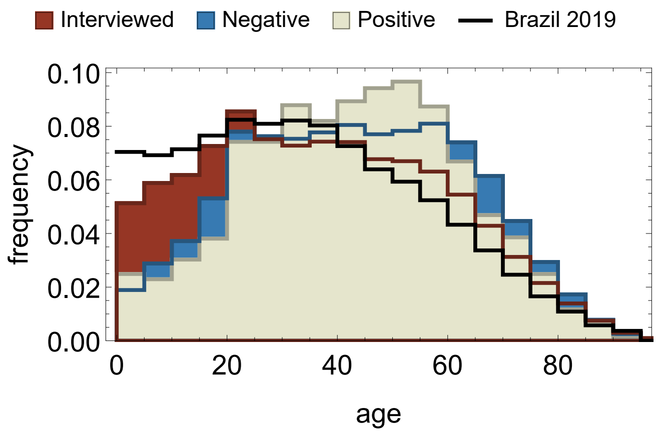


Figure S8. **Age distributions relative to the EPICOVID19-BR survey.** We split according to positive and negative test results and show also the age demographics of the Brazilian population and of the totality of interviewees in the survey.

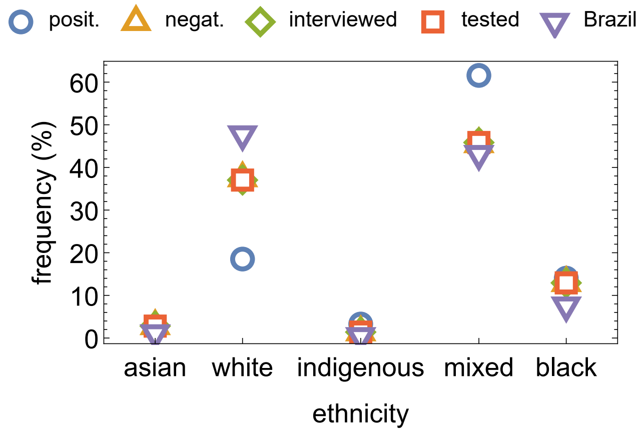


Figure S9. **Ethnic distribution in Brazil and in the EPICOVID19-BR survey.** We again split according to positive and negative test results and among all those interviewed and those who consented to be tested.

ethnicity.

### S7. FULL NUMERICAL RESULTS

Table S1 shows the full numerical results for $p_a$, $p_d$ and IFR for all states and all rounds of the EPICOVID19-BR survey. The values of IFR are marginalized over $T$, while the values of $p_d$ are relative to $T = 60$ days.

| State | $p_a$ R1 (%) | $p_a$ R2 (%) | $p_a$ R3 (%) | $p_d$ R1 (‰) | $p_d$ R2 (‰) | $p_d$ R3 (‰) | IFR R1 (%) | IFR R2 (%) | IFR R3 (%) | IFR all rounds (%) |
|---|---|---|---|---|---|---|---|---|---|---|
| AC | 3.9 (1.7–6.9) | 5.7 (3.4–8.6) | 4.9 (2.7–7.7) | 0.35 (0.32–0.37) | 0.53 (0.51–0.54) | 0.58 (0.58–0.58) | 0.72 (0.4–1.6) | 0.81 (0.53–1.4) | 1. (0.58–1.9) | 0.85 (0.63–1.2) |
| AL | 0.53 (0.–2.4) | 9.6 (6.2–14.) | 13. (9.3–18.) | 0.26 (0.24–0.28) | 0.43 (0.42–0.45) | 0.49 (0.48–0.49) | 1.6 (0.48–25.) | 0.41 (0.29–0.64) | 0.35 (0.23–0.51) | 0.47 (0.37–0.61) |
| AM | 11. (7.6–16.) | 13. (9.4–18.) | 7.5 (4.4–11.) | 0.64 (0.62–0.66) | 0.69 (0.69–0.69) | 0.55 (0.53–0.58) | 0.51 (0.36–0.77) | 0.48 (0.29–0.71) | 0.47 (0.28–1.3) | 0.5 (0.38–0.65) |
| AP | 8.6 (5.1–13.) | 14. (9.3–19.) | 13. (9.–18.) | 0.3 (0.27–0.34) | 0.43 (0.41–0.45) | 0.49 (0.47–0.5) | 0.32 (0.21–0.53) | 0.29 (0.2–0.43) | 0.34 (0.21–0.52) | 0.31 (0.24–0.4) |
| BA | 0.88 (0.16–1.8) | 3.9 (1.9–6.7) | 2.3 (0.91–4.2) | 0.12 (0.11–0.13) | 0.22 (0.21–0.23) | 0.3 (0.3–0.31) | 0.99 (0.47–3.9) | 0.47 (0.27–0.95) | 1. (0.54–2.5) | 0.81 (0.57–1.2) |
| CE | 6.7 (3.8–10.) | 13. (9.2–17.) | 18. (13.–22.) | 0.74 (0.71–0.78) | 0.96 (0.95–0.96) | 0.89 (0.87–0.91) | 0.98 (0.63–1.7) | 0.68 (0.47–1.) | 0.52 (0.29–0.7) | 0.67 (0.55–0.82) |
| DF | 0. (0.–1.5) | 0. (0.–2.2) | 0. (0.–2.2) | 0.068 (0.062–0.075) | 0.16 (0.14–0.17) | 0.28 (0.26–0.29) | 0.69 (0.13–27.) | 1.1 (0.25–31.) | 1.9 (0.44–54.) | 1.3 (0.61–3.5) |
| ES | 1.1 (0.–2.8) | 1.5 (0.43–2.8) | 2.6 (1.3–4.3) | 0.18 (0.17–0.2) | 0.37 (0.35–0.39) | 0.5 (0.49–0.51) | 0.92 (0.34–7.) | 1.9 (0.95–5.6) | 1.6 (0.94–3.2) | 1.6 (1.1–2.5) |
| GO | 0.3 (0.–1.2) | 0.38 (0.–1.2) | 0.95 (0.02–2.3) | 0.034 (0.032–0.036) | 0.068 (0.061–0.076) | 0.15 (0.14–0.16) | 0.39 (0.13–5.9) | 0.82 (0.29–9.9) | 0.96 (0.39–5.9) | 0.78 (0.47–1.6) |
| MA | 1.1 (0.–3.6) | 6.7 (4.4–9.4) | 7.1 (4.9–9.6) | 0.4 (0.39–0.42) | 0.53 (0.52–0.53) | 0.54 (0.54–0.54) | 1.6 (0.54–16.) | 0.71 (0.45–1.1) | 0.7 (0.43–1.1) | 0.82 (0.61–1.1) |
| MG | 0.95 (0.23–1.9) | 0.46 (0.08–0.92) | 0.63 (0.1–1.3) | 0.024 (0.023–0.026) | 0.049 (0.045–0.053) | 0.097 (0.09–0.1) | 0.18 (0.09–0.61) | 0.77 (0.37–3.2) | 1.1 (0.51–4.6) | 0.78 (0.52–1.3) |
| MS | 0. (0.–2.4) | 0.2 (0.–1.4) | 0.49 (0.–1.5) | 0.0074 (0.0069–0.0079) | 0.021 (0.018–0.025) | 0.068 (0.052–0.083) | 0.04 (0.01–1.2) | 0.22 (0.06–4.3) | 0.64 (0.23–7.2) | 0.4 (0.22–0.89) |
| MT | 4.3 (0.12–13.) | 0.86 (0.04–2.) | 0.61 (0.–1.5) | 0.035 (0.029–0.041) | 0.15 (0.13–0.17) | 0.38 (0.35–0.41) | 0.04 (0.01–0.26) | 1.1 (0.48–6.3) | 3.7 (1.5–26.) | 2.3 (1.4–4.4) |
| PA | 12. (9.1–15.) | 13. (10.–16.) | 6.3 (4.5–8.4) | 0.7 (0.67–0.74) | 0.9 (0.9–0.91) | 0.83 (0.81–0.86) | 0.56 (0.44–0.75) | 0.66 (0.5–0.87) | 1.3 (0.63–1.9) | 0.66 (0.56–0.78) |
| PB | 1.5 (0.05–3.8) | 4.8 (2.9–7.3) | 1.7 (0.45–3.3) | 0.15 (0.14–0.16) | 0.28 (0.26–0.29) | 0.42 (0.4–0.43) | 0.57 (0.22–3.4) | 0.51 (0.34–0.86) | 1.8 (0.88–5.8) | 0.89 (0.64–1.3) |
| PE | 2.4 (0.76–4.8) | 1.8 (0.34–4.) | 0.64 (0.–2.) | 0.48 (0.45–0.51) | 0.67 (0.66–0.68) | 0.71 (0.7–0.71) | 1.5 (0.74–4.2) | 2.5 (1.1–9.7) | 5. (1.5–54.) | 2.7 (1.7–4.8) |
| PI | 3. (0.4–7.5) | 1.3 (0.28–2.7) | 8.1 (5.4–11.) | 0.12 (0.1–0.13) | 0.3 (0.28–0.32) | 0.49 (0.47–0.51) | 0.24 (0.1–1.1) | 1.7 (0.79–6.) | 0.56 (0.39–0.84) | 0.78 (0.58–1.1) |
| PR | 0.53 (0.02–1.2) | 0.64 (0.–1.8) | 0.79 (0.21–1.5) | 0.025 (0.023–0.027) | 0.052 (0.048–0.056) | 0.1 (0.092–0.11) | 0.29 (0.12–1.7) | 0.42 (0.15–3.9) | 0.97 (0.49–3.1) | 0.69 (0.45–1.2) |
| RJ | 1.6 (0.11–3.8) | 5.5 (2.9–9.) | 7.9 (4.8–12.) | 0.51 (0.48–0.54) | 0.72 (0.71–0.74) | 0.79 (0.78–0.79) | 2. (0.8–11.) | 1.1 (0.67–2.1) | 0.87 (0.5–1.5) | 1.2 (0.85–1.6) |
| RN | 2. (0.36–4.4) | 3.2 (1.3–5.7) | 4.9 (2.7–7.7) | 0.15 (0.14–0.17) | 0.33 (0.3–0.36) | 0.55 (0.53–0.57) | 0.52 (0.23–2.) | 0.84 (0.46–2.) | 0.98 (0.61–1.8) | 0.85 (0.61–1.3) |
| RO | 0.05 (0.–2.2) | 1.9 (0.32–4.3) | 5.3 (2.8–8.6) | 0.25 (0.22–0.27) | 0.48 (0.47–0.5) | 0.6 (0.59–0.61) | 1.7 (0.43–40.) | 1.7 (0.73–6.8) | 0.95 (0.56–1.8) | 1.3 (0.85–2.) |
| RR | 3.4 (1.2–6.7) | 24. (18.–29.) | 20. (15.–26.) | 0.24 (0.23–0.26) | 0.46 (0.43–0.48) | 0.67 (0.65–0.68) | 0.55 (0.28–1.5) | 0.19 (0.15–0.24) | 0.31 (0.22–0.42) | 0.26 (0.22–0.32) |
| RS | 0.43 (0.01–0.93) | 0.45 (0.01–0.99) | 0.46 (0.04–0.98) | 0.025 (0.024–0.026) | 0.038 (0.036–0.04) | 0.066 (0.062–0.071) | 0.37 (0.16–2.3) | 0.54 (0.22–3.3) | 0.97 (0.42–5.1) | 0.67 (0.43–1.2) |
| SC | 0.5 (0.09–0.99) | 0.51 (0.1–0.99) | 0.59 (0.13–1.1) | 0.022 (0.021–0.022) | 0.031 (0.029–0.033) | 0.054 (0.049–0.058) | 0.31 (0.15–1.2) | 0.44 (0.21–1.7) | 0.68 (0.32–2.4) | 0.49 (0.33–0.82) |
| SE | 0. (0.–1.6) | 0.33 (0.–2.) | 2.7 (0.75–5.5) | 0.11 (0.1–0.13) | 0.29 (0.27–0.32) | 0.52 (0.5–0.55) | 1.1 (0.27–29.) | 2.1 (0.62–39.) | 1.4 (0.68–4.4) | 1.5 (0.92–3.) |
| SP | 2. (0.41–4.3) | 1.1 (0.–2.8) | 0.69 (0.–2.1) | 0.27 (0.26–0.29) | 0.36 (0.35–0.36) | 0.4 (0.39–0.4) | 0.93 (0.42–3.4) | 1.8 (0.65–12.) | 2.7 (0.88–28.) | 1.7 (1.1–3.3) |
| TO | 0.35 (0.–1.1) | 0.58 (0.–1.5) | 1.2 (0.35–2.3) | 0.058 (0.053–0.063) | 0.12 (0.11–0.12) | 0.16 (0.16–0.17) | 0.78 (0.29–9.1) | 1.1 (0.43–9.2) | 0.99 (0.48–3.) | 0.97 (0.63–1.7) |

Table S1. Maxima of the probability distributions and 95% CI. IFR results are marginalized over $T$, whereas $p_d$ is relative to $T = 60$ days and given in parts per thousands.
